# Monoclonal antibody levels and protection from COVID-19

**DOI:** 10.1101/2022.11.22.22282199

**Authors:** Eva Stadler, Martin T Burgess, Timothy E Schlub, Khai Li Chai, Zoe K McQuilten, Erica M Wood, Mark N Polizzotto, Stephen J Kent, Deborah Cromer, Miles P Davenport, David S Khoury

## Abstract

Multiple monoclonal antibodies have been shown to be effective for both prophylaxis and therapy for SARS-CoV-2 infection. Here we aggregate data from randomized controlled trials assessing the use of monoclonal antibodies in preventing symptomatic SARS-CoV-2 infection. We use data on changes in the *in vivo* concentration of monoclonal antibodies, and the associated protection from COVID-19, over time to model the dose-response relationship of monoclonal antibodies for prophylaxis. We estimate that 50% protection from COVID-19 is achieved with a monoclonal antibody concentration of 54-fold of the *in vitro* IC50 (95% CI: 16 – 183). This relationship provides a quantitative tool allowing prediction of the prophylactic efficacy and duration of protection for new monoclonal antibodies administered at different doses and against different SARS-CoV-2 variants.

Finally, we compare the relationship between neutralization titer and protection from COVID-19 after either monoclonal antibody treatment or vaccination. We find no evidence for a difference between the 50% protective titer for monoclonal antibodies and vaccination.

## Introduction

Vaccination has been shown to be highly effective at preventing both symptomatic and severe COVID-19 (reviewed by Cromer et al.^1^). However, vaccination is less effective in many immune compromised and elderly individuals where immunogenicity and clinical data show considerably impaired responses to vaccination^2,3^. Multiple monoclonal antibody products have been shown to be effective as pre- and post-exposure prophylaxis against pre-Omicron variants^4-6^, as well as when administered therapeutically early in infection^7-11^. We recently analysed the available data on antibody treatment of symptomatic SARS-CoV-2 infection to determine the dose-response relationship between the antibody dose administered (after conversion to a neutralizing dose equivalence) and the protection from progression to hospitalisation^12^. However, the dose-response curve for monoclonal antibody administration as prophylaxis of COVID-19 has not yet been determined, since there have to-date been fewer randomized control trials assessing the prophylactic effect. Here we adopt an alternative approach, comparing the loss of antibody *in vivo* with the loss of efficacy of monoclonal antibodies over time following administration. In addition, we use data on the loss of neutralisation and protection observed to new variants to inform this relationship^13^. Using this data on temporal changes in monoclonal antibody concentration and changes in potency to new variants, we estimate the relationship between *in vivo* antibody concentration and protection, which may provide a valuable clinical tool for predicting the efficacy of new monoclonal products and existing products against new variants^12^. Finally, we assess whether neutralizing antibodies mediate protection or merely correlate with protection by comparing the relationship between neutralization titer and protection after vaccination^14^ and in naïve individuals receiving monoclonal antibodies.

Together this work provides a quantitative framework for dissecting the mechanisms of protection in vaccination and informing the use of critical immunotherapies.

## Results

### Aggregating studies of monoclonal antibodies as prophylaxis

We searched MEDLINE, PubMed, Embase and the Cochrane COVID-19 Study Register for randomized placebo-control trials of SARS-CoV-2-specific monoclonal antibodies (mAbs) used as pre-exposure and peri-exposure prophylaxis for COVID-19. We included only studies where both protection from symptomatic infection and pharmacokinetic information of the monoclonal antibody were provided within the same study. We identified six eligible studies assessing monoclonal antibodies as pre-exposure and peri-exposure prophylaxis for COVID-19^4-6,15,16^ (see Table S1). The antibodies used in these studies were casirivimab/imdevimab (3 studies), bamlanivimab, tixagevimab/cilgavimab, and adintrevimab. One of these studies did not provide data on the pharmacokinetics of the antibody (bamlanivimab)^16^ and was excluded. Of the remaining four studies, three reported a break-down of cases in treatment and control arms by time since administration and two studies had data on the timing of cases that could be extracted from the publication^4-6,15^. Four of these five studies assessed protection at a time before the omicron variants were the dominant circulating variants and against variants where the antibody had been shown to be able to neutralize the virus *in vitro*^17^. One study assessed protection in two time periods; firstly in a pre-Omicron period when the Delta variant was the dominant circulating variant, and separately later when Omicron variants BA.1 and BA.1.1 were the dominant variants^13^. The overall efficacies against pre-Omicron variants in the included studies ranged from between 66.7 – 92.4%. We identified a trend for lower efficacies with increasing time since administration and against the escaped variant (Figure 1).

**Figure 1:**
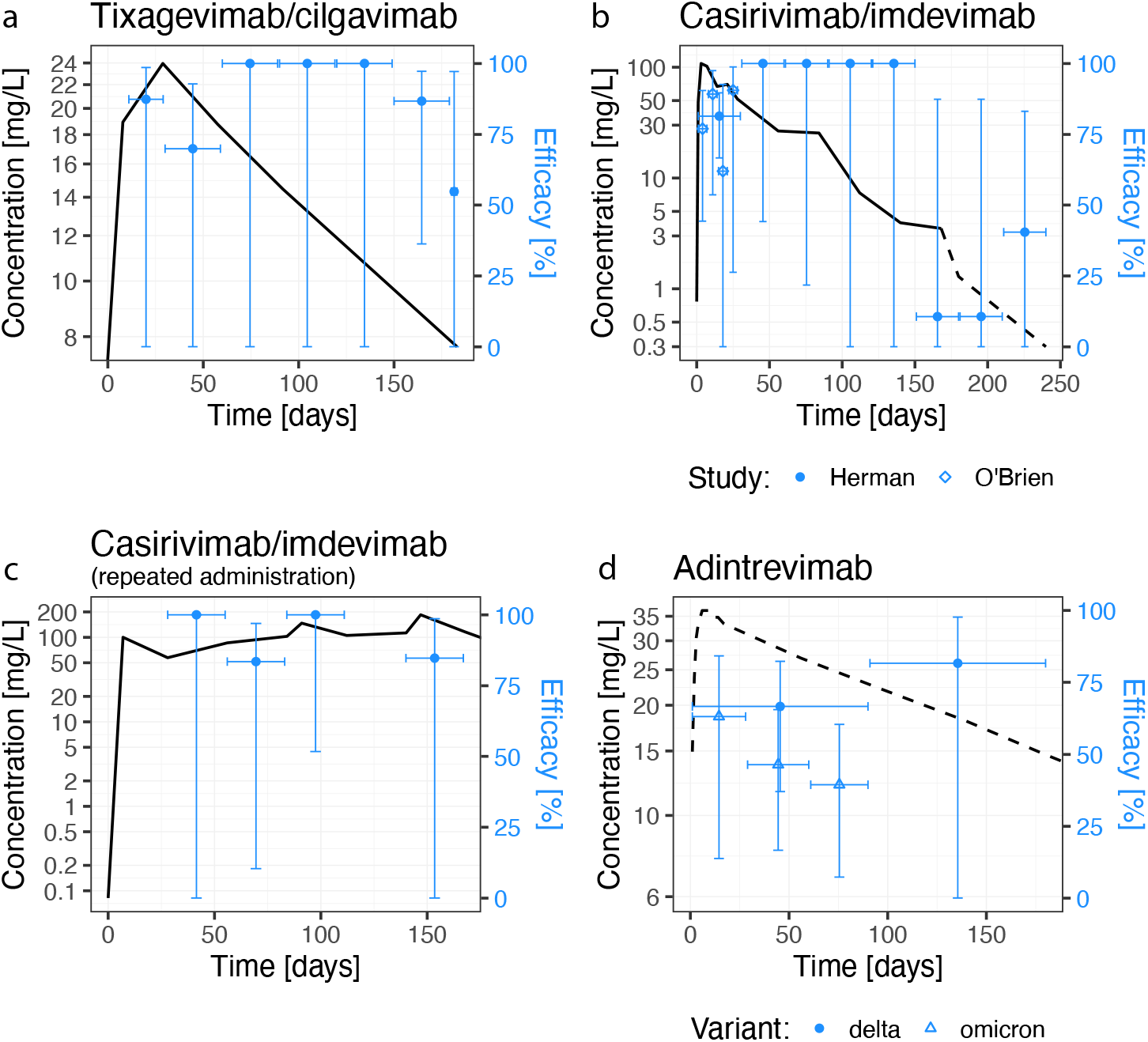
Reported protection and antibody concentration from three RCTs of monoclonal antibodies in preventing COVID-19. The efficacy at each time interval is shown in blue (horizontal error bars indicate time interval and vertical error bars represent 95% CI). The antibody concentration is shown in black. (a) Antibody concentration data for tixagevimab/cilgavimab was extracted from Levin et al. (b) Single administration of casirivimab/imdevimab data is a combination of data from O’Brien et al. and Herman et al. who report on the same clinical trial over different follow-up intervals. Efficacy data was reported frequency over the first four weeks in O’Brien et al. (diamonds), and monthly for eight months in Herman et al. (circles). Antibody concentration data was reported up to day 168 in O’Brien et al. (solid line, panel b), and modelling of the pharmacokinetic profile of the antibody concentration, reported in Herman et al., was used to inform the antibody concentration between 168 and 240 days (dashed line, panel b). (c) Isa et al. reported efficacy and *in vivo* concentration after repeated administration of 1.2g of casirivimab/imdevimab every four weeks. Hence, the antibody concentration did not decline as in the other studies. (d) The modelled concentration of adintrevimab after a single administration was extracted from the study by Schmidt et al. The efficacy of adintrevimab was reported both when the delta variant was dominant (circles) and when omicron variants BA.1 and BA.1.1 were dominant (triangles).

### A significant dose-response relationship between protection and mAb concentration

To investigate whether declining efficacy with time and new variants was indicative of a dose-response relationship between mAb concentration and efficacy, we compared the antibody concentrations reported within different time intervals in each study with the reported efficacy at the corresponding time interval (Figure 2). To compare between antibodies of different potencies and against different variants, we normalized antibody concentration using the *in vitro* IC50 for each antibody/variant combination from a meta-analysis of *in vitro* studies (Table S2 and Figure S1, as previously reported^12^ using data from Tao et al.^18^). We found a significant relationship between efficacy and mAb concentration (as a fold of the *in vitro* IC50) (p<0.0001, generalized linear mixed model (GLMM) and chi-squared test, see Methods). In a leave-one-out analysis, we found this relationship remained significant when any one study is omitted (Table S3). Fitting a logistic dose-response relationship to this data, we estimated a peak efficacy of 94% (95% CI: 84 – 106%), and concentration for 50% of efficacy of 54-fold *in vitro* IC50 (95% CI: 16 – 183) (Table S4). Against the ancestral virus, this equates to a plasma antibody concentration of 0.22 mg/L concentration for tixagevimab/cilgavimab, 0.18 mg/L for casirivimab/imdevimab and 0.36 mg/L for adintrevimab being required for 50% protection against COVID-19.

**Figure 2:**
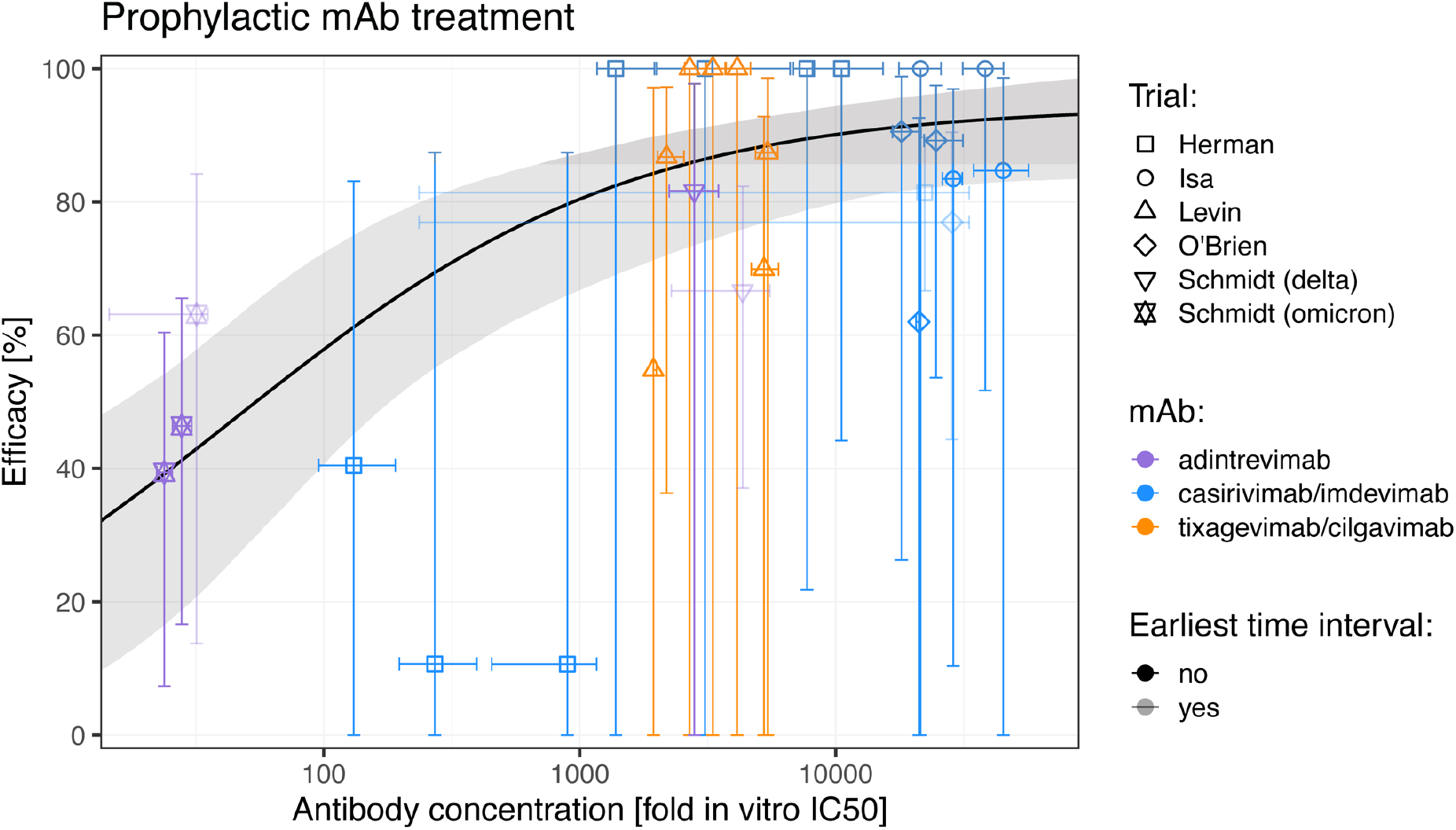
Dose-response relationship between antibody concentration and protection. The estimated geometric mean antibody concentration and protective efficacy in the matching study and time interval (expressed as a fold of the *in vitro* IC50 of each antibody) is shown. Horizontal error bars indicate the range of the (means of the) antibody concentrations reported during each time interval, and vertical error bars indicate the 95% confidence interval of the efficacy. We estimate a dose-response relationship (black line) by fitting a logistic model to the data and estimate the 95% confidence intervals using parametric bootstrapping (grey shading). Efficacy data reported early (in the first 7, 10, 28 or 90 days, depending on the first time point reported in the study) after treatment were excluded from the model fitting (low opacity data points), as antibody concentration changed rapidly over this time interval and ensured exclusion of incidental treatments which occurred after an unidentified infection.

### Predicting monoclonal antibody efficacy against new variants

A major challenge in the COVID-19 pandemic has been in decision-making around whether pharmaceuticals shown to be effective against ancestral SARS-CoV-2 should continue to be used when new variants emerge. This is especially true for monoclonal antibody therapies, where recommendations on the use of mAb therapeutics have changed numerous times with the emergence of each Omicron subvariant^19,20^. This has been particularly difficult when a mAb loses partial, but not complete, recognition of a new variant. If we assume that the relationship defined here between antibody concentration (normalised to *in vitro* neutralising IC50) and efficacy will continue to hold for different variants of concern, as it has for vaccine-induced neutralizing antibodies^1,21^, we can use the dose-response relationship in Figure 2 to estimate the loss of efficacy and duration of protection of monoclonal antibodies to new variants. For example, tixagevimab/cilgavimab administered intramuscular at a dose of 300 mg is predicted to maintain >50% protection for 670 days (95% CI: 503 – 836 days) against the ancestral variant where the *in vitro* IC50 is 4.00 ng/mL and the half-life of antibody 95 days (Figure 3, Tables S2, S5 and S6). However, if the *in vitro* IC50 against a new variant were increased (but still detectable), as is the case for some mAbs against the Omicron subvariants (Table S2), the model allows us to predict the reduction in the time above 50% protection. In the meta-analysis used here on mAb IC50’s against different variants, tixagevimab/cilgavimab was found to maintain detectable *in vitro* neutralization titers against the BA.1, BA.2 and BA.4/5 Omicron subvariants (see Table S2, meta-analysis as described in^12^, using data from Tao et al.^18^). The increase in IC50 for this antibody combination against is predicted to reduce efficacy at the peak antibody concentration from 88.8% (95% CI: 77.3 – 93.0) against ancestral virus, to 56.7% (95% CI: 38.9 – 71.4), 77.0% (95% CI: 62.8 – 85.4) and 43.4% (95% CI: 21.1 – 58.5) efficacy against the BA.1, BA.2 and BA.4/5 subvariants, respectively (Table S7). Using the dose-response relationship here (Figure 2), this would predict that the duration of >50% protection of tixagevimab/cilgavimab against BA.1 and BA.2 will be 97 (95% CI: 0 – 263) and 360 (95% CI: 194 – 526) days, respectively (Table S5). On the other hand, casirivimab/imdevimab and adintrevimab have larger increases in the IC50 against these subvariants (Table S2) and are not predicted to maintain >50% efficacy to these variants (Tables S5 and S7).

**Figure 3:**
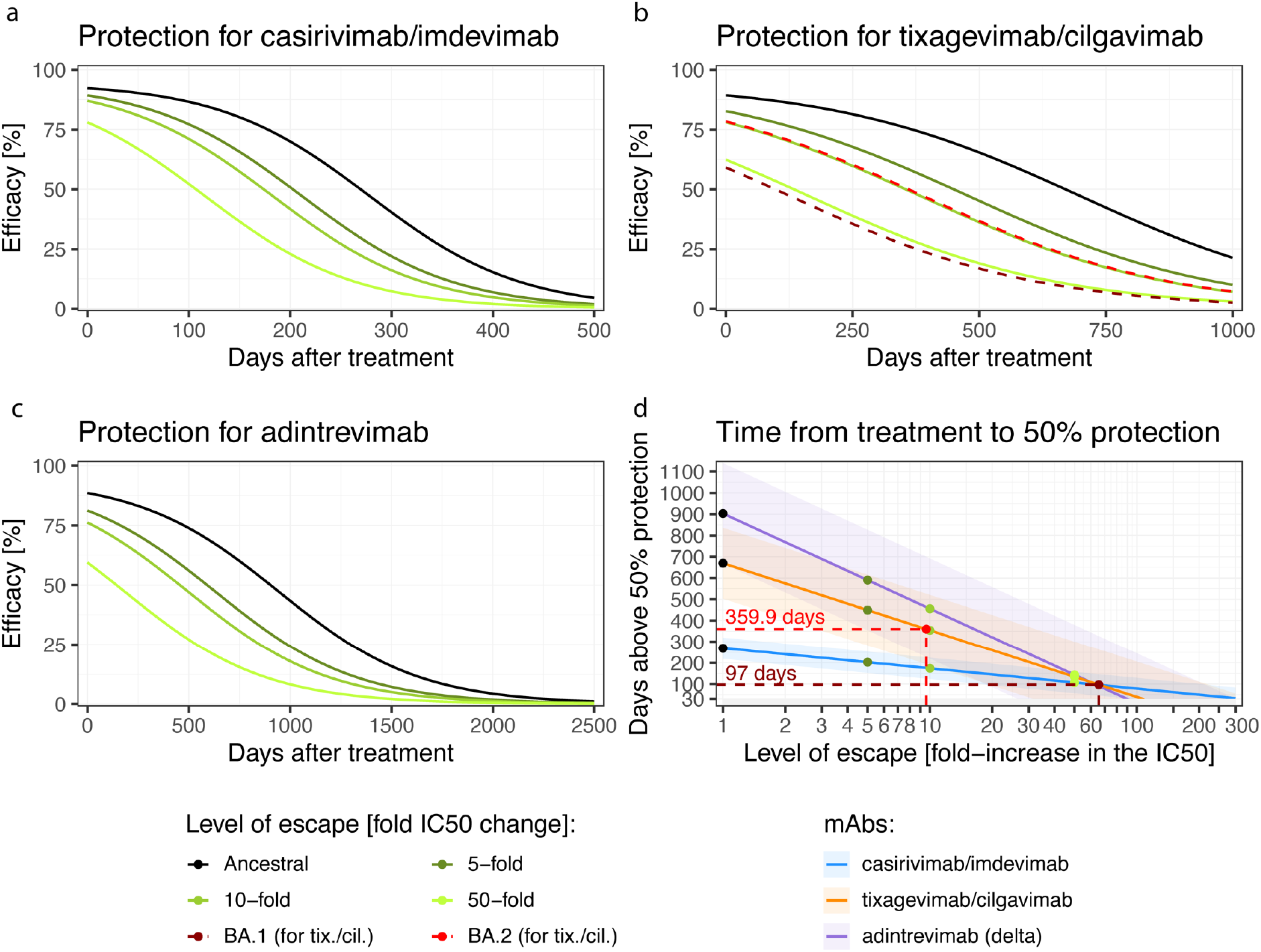
Duration of protection against SARS-CoV-2 variants. Using the dose-response relationship of antibody concentration and protective efficacy in Figure 2 and the estimated half-life of antibodies after treatment (28.8 days (95% CI: 26.6 – 31.3) for casirivimab/imdevimab, 94.9 days (95% CI: 84.2 – 108.9) for tixagevimab/cilgavimab and 134.7 days (95% CI: 132.7 – 136.7) for adintrevimab, see Figure S2 and Table S5), we predict the efficacy over time for these antibody combinations (black line, a, b and c). We also estimate the protection over time curves of these antibody combinations given different fold-increases in the IC50 that may be experienced to new variants (coloured lines). (d) For each hypothetical loss of potency of these antibodies (i.e. fold increase in IC50, x-axis), we predict the number of days each antibody will remain above 50% protection (i.e. the number of days the mean antibody concentration will remain above the level associated with 50% protection of 54-fold-IC50, Figure 2). The shaded regions indicate the 95% confidence interval of the duration of protection (using the 95% CI for 50% protection: 16 – 183). We note that the casirivimab/imdevimab and the tixagevimab/cilgavimab combinations and adintrevimab are expected to maintain more than 50% protection for approximate 269, 670 and 903 days against a variant with the same IC50 as the ancestral virus, respectively, and can tolerate up to a 319.6-, 106.8- and 89.6-fold drop in potency to a new variant, respectively, and still be expected to maintain 30 days of more than 50% protective efficacy. The increase in IC50 against BA.4/5 is sufficient to reduce efficacy below 50% for all three products.

Another formulation of this question is to ask “What is the maximum increase of IC50 (drop in neutralization titer) that can be tolerated while still maintaining a minimum duration of protection?”. For example, if we wish to provide a period of at least 30 days with >50% protection, then tixagevimab/cilgavimab, casirivimab/imdevimab and adintrevimab, at the current doses, can tolerate at most 106.8-fold (95% CI: 31.7 – 360.0), 319.5-fold (95% CI: 94.8 – 1076.5) and 89.6-fold (95% CI: 26.6 – 302.0) increases in *in vitro* IC50, respectively (Table S8). Figure 3d shows the predicted duration of >50% protection for casirivimab/imdevimab, tixagevimab/cilgavimab and adintrevimab for any given change in *in vitro* IC50. This analysis provides a tool to determine how regularly monoclonal antibodies may need to be re-administered to provide high confidence that subjects will maintain a serum antibody concentration that will provide more than 50% efficacy against the circulating variant.

### Comparing monoclonal antibody prophylaxis with vaccine-induced protection

Multiple lines of evidence have established that neutralizing antibody titers correlate with protection from COVID-19 in vaccinated individuals^14,22-24^. An important question in whether neutralizing antibodies are mechanistic in mediating this protection, or merely correlate with protection^25^. Similarly, if antibodies are able to directly mediate protection, identifying the magnitude of their contribution to overall protection (compared to other mechanisms) is an important question. One way to address this is to compare the level of protection achieved after administration of antibodies alone with that achieved after vaccination.

Antibody administration alone should reflect the antibody-related contribution to protection, while vaccination should incorporate both antibody- and cell-mediated protection. Recently, Schmidt et al., reported the loss of *in vitro* neutralisation of adintrevimab to Omicron BA.1 and BA.1.1 resulted in a corresponding loss of efficacy that was not inconsistent with the relationship between reports of vaccine effectiveness^13^. Here we address this question by integrating the available data across currently available studies on monoclonal antibody prophylaxis. We then used an established correlate of vaccine protection for COVID-19 to analyse whether prophylaxis against COVID-19 after passive antibody administration is achieved at similar levels of neutralisation to protection observed after vaccination (Figure S3).

Figure 4a compares the efficacy of high-potency mRNA vaccines^26,27^ and monoclonal antibody prophylaxis in prevention of symptomatic SARS-CoV-2 infection with the ancestral or delta variant. We find that the observed mean protection achieved with mAbs of 84.8% (95% CI: 76.0 – 90.8) was significantly different to the efficacy achieved with vaccination of 94.5% (95% CI: 91.6 – 96.7) (p=0.002, GLMM, see Methods). However, a similar efficacy may be possible by administering monoclonal antibodies at much higher neutralizing antibody titers compared with the neutralising antibody titers achieved in vaccinated individuals.

**Figure 4:**
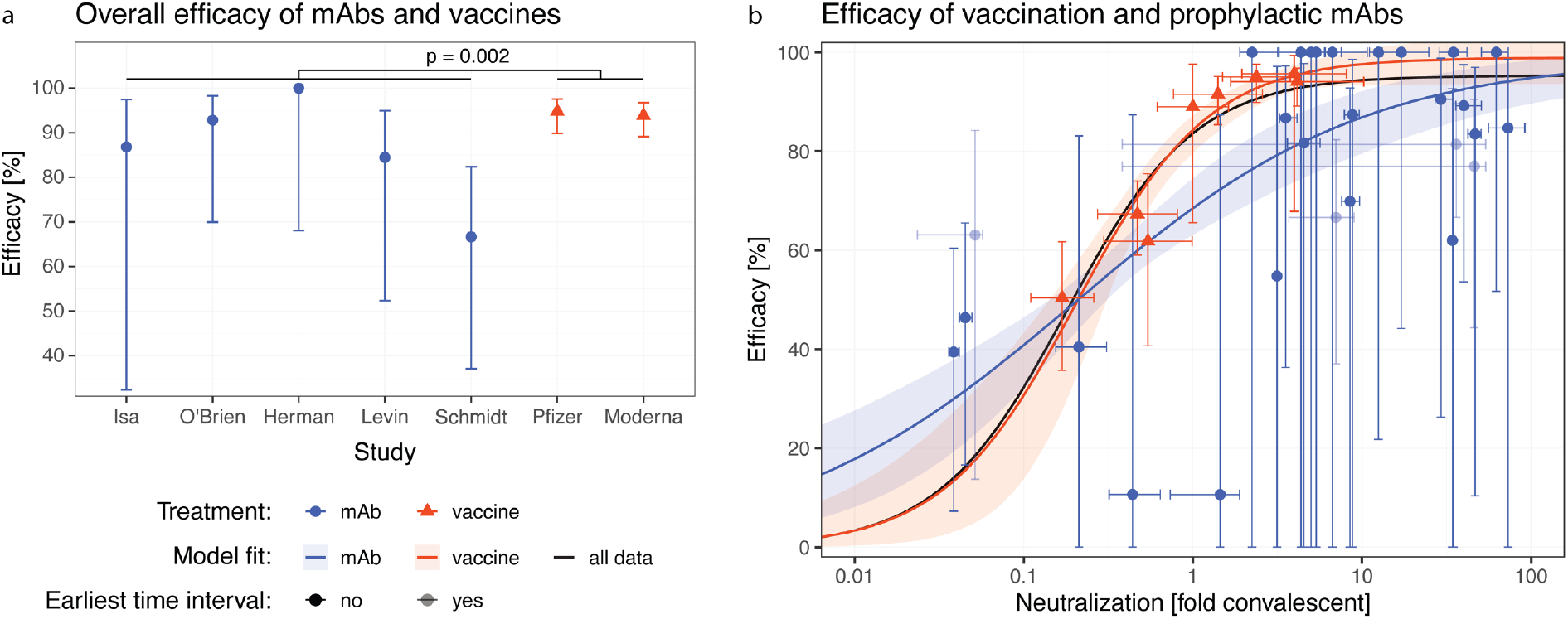
Comparison of dose response curves after vaccination and monoclonal antibody administration. (a) We compared the overall efficacies of prophylactic mAb treatment and high-potency RNA vaccines and found a significantly difference in efficacy (p = 0.002, GLMM, see Methods). The mean efficacy in the mAb studies was 84.8% (95% CI: 76.0 – 90.8%) and for the high-potency mRNA vaccines the mean efficacy was 94.5% (95% CI: 91.6 – 96.7%). (b) We normalized the neutralization titers to a common scale of ‘fold-of-convalescent’ titer to allow direct comparison taking differences in the neutralization into account. We then fitted the combined data with a logistic model in which the same three parameters were used for maximum efficacy, neutralization for 50% efficacy, and slope (b, black line). Next, we allowed parameters to vary between the monoclonal and vaccination data fitting. Here we show the best fit to the data was with a model where the slope is allowed to vary between mAb studies (blue) and vaccine studies (red) but the maximal efficacy and the neutralization giving 50% protection were the same for both types for treatment (based on model comparisons with the likelihood-ratio test, see Figure S3).

Thus, we next compared the level of protection achieved for a given neutralizing antibody titer after either vaccination (from Khoury et al.^14^) or after treatment with monoclonal antibodies (Figure 4b). To compare titers between vaccination and monoclonal antibody administration, we normalized the titers of each to a scale relative to the geometric mean titer of neutralizing antibodies seen in convalescent individuals against ancestral virus, after the first wave of COVID-19 (a ‘fold-convalescent’ scale, as previously described for the studies of vaccination^14^, and as described in the Methods for mAbs). In order to test whether the neutralizing antibody titer associated with a given level of protection for vaccines and mAbs were consistent or different, we fitted these data together using the same parameters for the monoclonal antibody and vaccine dose-response relationships. We then tested whether the model fitted better with separate parameters for mAb treatment and vaccination (likelihood ratio test, see Methods and Figure S3). There was no evidence of a difference in the neutralisation titre required for 50% protection (p = 0.48, Figure S4).

However, this analysis showed that the best-fit model was one where the same dose-response relationship existed for both vaccination and mAbs but with the estimated slope being higher for vaccination. Alternatively, a model with the same parameters except a different maximum efficacy had a nearly equivalently good fit (AIC difference 1.9, Table S9). In these two nearly equivalently good models, at neutralisation titres above the 50% protective level, the efficacy of mAb tended lower than that achieved by vaccination (Figure 4b, Figure S3, Table S9). Together, these results indicate that similar levels of neutralising antibodies from either vaccination or the administration of monoclonal antibody are associated with 50% protection from COVID-19, but with a trend towards a lower maximum protection from COVID-19 that can be achieved with monoclonal antibodies compared to vaccination.

## Discussion

Here we demonstrate a relationship between the monoclonal antibody concentration and efficacy to prevent COVID-19. Further, we estimate the concentration of antibody required to have a high confidence of maintaining at least 50% protection. Our model fitting enabled us to quantify the uncertainty in this relationship and estimate that if a treated population can maintain a mean *in vivo* monoclonal antibody concentration of > 54-fold (95% CI: 16 – 183) of the *in vitro* IC50 of the antibody to the circulating variant, they should maintain > 50% efficacy against COVID-19. Analysis of the dose-response curve for monoclonal antibodies allows prediction of the level and duration of protection against different SARS-CoV-2 variants (Figure 3). Our results suggest that casirivimab/imdevimab, tixagevimab/cilgavimab and adintrevimab would provide >50% protection against the ancestral SARS-CoV-2 strain for 269, 670 and 903 days respectively (Table S5). At the current doses, casirivimab/imdevimab and adintrevimab are not predicted to achieve 50% protection against the Omicron BA.2 variant (Table S7). For tixagevimab/cilgavimab, the protective interval against the BA.2 variant is dramatically reduced from 670 days to 360 days, and none of these antibodies were predicted to achieve 50% protection against the Omicron BA.4/5 variant (Tables S5 and S7). Counterintuitively, although antibodies with a longer half-life are expected in general to provide protection for longer, these are also expected to lose more ‘days of protection’ for a given fold-increase in IC50 (to a new variant), compared to mAbs with shorter half-lives (Figure 3). The higher susceptibility of therapeutics with longer half-lives to fold-shifts in the IC50 has been discussed previously for antimalarial products^28^, and can be explained by considering that when antibodies lose 2-fold neutralization against a new variant it is equivalent to the mAb losing one half-life of time above a threshold. Therefore, a mAb with a 100-day half-life will lose 100 days above a specified threshold, whereas a mAb with only a 30-day half-life will lose 30 days above the same threshold.

The estimated *in vivo* concentration of antibody required for 50% protection from COVID-19 is much higher than the level of antibody required to neutralize virus *in vitro* (approx. 50-fold), suggesting that *in vivo* neutralization may be much less efficient than the observed neutralization *in vitro*. This difference between *in vitro* IC50 and the *in vivo* 50% protective titer is not unexpected, given the major differences between infection in these environments. For example, *in vitro* neutralization assays usually involve pre-incubation of antibody and virus for an hour before exposure to cells. Similarly, the *in vitro* IC50 in plaque reduction assays estimates the antibody concentration required to neutralize 50% of virions. However, the dose required to completely neutralize large inocula may be considerably higher^29^. In addition, *in vivo* antibody titers are assessed in the serum. However, antibody concentration at the mucosa is lower than the plasma level^30^, and thus higher (serum) titers may be required to achieve neutralization on mucosal surfaces.

We and others have previously shown that neutralizing antibodies are a correlate of protection from COVID-19^14,22,31-33^. A major question in understanding vaccine-mediated immunity is whether neutralizing antibodies are simply a surrogate marker of protection or are mechanistic in protecting individuals from symptomatic infection^14^. To-date it has only been possible to consider this question indirectly. For example, we have noted that the drop in neutralizing antibodies against new variants and over time both provide good predictions of the change in efficacy of vaccines over time and against new variants^1,34^. Schmidt et al. recently showed that the monoclonal antibody adintrevimab lost neutralisation to Omicron BA.1 and BA.1.1, and a corresponding loss of prophylactic efficacy compared with that predicted by the relationship between neutralisation titre and vaccine effectiveness studies^13^. Here, we extracted data on antibody PK and temporal changes in efficacy from five monoclonal antibody studies (including from Schmidt et al.^13^), and compared this with an established immune correlate of protection after vaccination that has been validated across a number of settings^1,14,21,35,36^. This allows a direct comparison of the protection provided by passively administered antibodies versus vaccine-induced. We have been able to show that the neutralization titer required for 50% protection by vaccination or monoclonal antibodies is comparable (Figure 4), although a predicted trend towards higher vaccine efficacy at high neutralising antibody titers was observed (Figure S4). This suggests that neutralizing antibodies are sufficient to explain much of the protection from symptomatic infection induced by vaccination.

The difference in protection at a given neutralization titer between vaccination and monoclonal antibody therapy may be due to the additional benefit in vaccinees of a polyclonal antibody response, other non-neutralizing functions of antibodies, recall of immune memory and/or other cellular immune responses. These functions may contribute to the estimated trend towards higher maximal protection for vaccines. Also, the efficacy studies of tixagevimab/cilgavimab and adintrevimab had infection events with other pre-Omicron variants, in particular the delta variant, and we have assumed that the *in vitro* IC50’s of these antibodies (especially) against the delta variant were equivalent to the IC50s against ancestral virus, for simplicity. However, it is likely that there was some increase in IC50 against the delta variant which we have not accounted for and this may explain some of the apparent lower efficacy in these studies (Figure 4a). While our analysis has shown that neutralizing antibodies are sufficient for protection from COVID-19, it is not possible to conclude from this analysis that neutralizing antibodies are necessary for protection. We note that evidence in animal models supports the findings that neutralising antibodies mediate protective immunity^37^, with some showing an additional benefit of Fc-receptor function^38^.

The analysis presented here has a number of limitations. Firstly, our dose-response analysis requires comparison of the *in vivo* measured antibody concentrations and the estimated *in vitro* IC50s. We also assume the population average half-life of antibodies in the treated population, rather than assessing individual titers over time. Further, we are reliant upon summary data on monoclonal antibody protection broken down by time as reported in, or extracted from, the published studies, and thus could not account fully for subjects lost to follow-up (although fortunately these numbers are relatively small). Additionally, the analysis is strongly influenced by the results from the Herman et al.^15^ and Schmidt et al.^13^ studies given their contribution of data at lower antibody concentrations (Figure 2). To gain more robust estimates of the dose-response curve for the use of monoclonal antibodies for prophylaxis, studies with longer follow-up times would be greatly beneficial where this is ethical.

Another factor affecting our analysis is the different mode of delivery of the monoclonal antibodies. Casirivimab/imdevimab was administered subcutaneously in the Isa^5^, O’Brien^6^ and Herman^15^ studies, whereas tixagevimab/cilgavimab and adintrevimab were administered intramuscularly^4,13^. Plasma antibody concentrations had much slower increases following intramuscular administration compared with subcutaneous administration of casirivimab/imdevimab, and thus it is possible that there is a delay until protective antibody concentrations are achieved (Figure 1). To avoid this difference and also to account for the risk of infection at the time of antibody administration, we omitted the earliest time interval from our analysis (which encompassed the first 7, 10, 28 or 90 days across different studies).

Studies of *in vitro* neutralization of different SARS-CoV-2 variants provide a range of different IC50’s for each antibody. We therefore used a meta-analysis of *in vitro* IC50’s to inform the mean IC50 for each antibody and to normalize antibody levels to the average convalescent titer for comparison with vaccine titers^18^. Only a subset of these studies reported the geometric mean neutralization titer of a cohort of convalescent plasma from early in the pandemic (n=13). This meta-analysis aggregated IC50’s estimated from a wide range of neutralization assays, and the definition of convalescent sera was specified differently in each study, introducing some potential confounders to these aggregated estimates. However, our multiple regression model included random effects for study, which can account for systematic differences in neutralization in the studies – for example due to assay differences.

## Conclusion

Vaccination has provided a high level of population immunity to COVID-19. However, there remain a number of subgroups in which vaccination is either not possible or is ineffective (largely due to immunodeficiency). The use of monoclonal antibodies for prophylaxis in these cohorts has the potential to provide long-term protection from both symptomatic and severe COVID-19 for these vulnerable groups^4-11,15,16^. However, the frequent observation of novel SARS-CoV-2 variants that escape antibody recognition has raised significant challenges in predicting monoclonal antibody protection against new variants. Further work is required to obtain more data on protection at low antibody levels, as well as to validate predictions of prophylactic efficacy against SARS-CoV-2 variants. Within this context, the work presented here provides a quantitative and evidence-based framework for predicting monoclonal antibody efficacy that can be used in the assessment of novel therapeutics or in designing optimal regimes for new SARS-CoV-2 variants.

## Methods

### Search strategy for studies of COVID-19 prophylaxis with monoclonal antibodies

We searched MEDLINE, PubMed, Embase and the Cochrane COVID-19 Study Register from inception to 30 November 2022, for randomized control trials of monoclonal antibodies for the prevention of COVID-19. We identified six studies. We included only the five studies where monoclonal antibody concentration was also reported for the cohort (Table S1). These studies were in a mixture of true pre-exposure prophylaxis and peri-exposure prophylaxis settings (Table S1).

Two studies, O’Brien et al.^6^ and Herman et al.^15^, reported results from the same clinical trial over different follow-up intervals (4 weeks and 8 months respectively). Thus, to avoid duplication of the same trial results, we integrated the results from these studies. In particular, the O’Brien trial reported outcomes on a weekly basis for 4 weeks whereas the Herman trial reported outcomes on a monthly basis for 8 months. Therefore, for these trials the weekly outcomes reported in O’Brien were used for the weeks 2 to 4 after administration (the initial week was omitted due to rising antibody levels in this period) and the results from Herman et al., were used for the months 2 to 8 only. In addition, antibody concentration data for the cohort was extracted from Figure S4 of O’Brien et al. from 0-168 days, whereas the Herman reported only pharmacokinetic model predictions of the concentration over the interval of 30-240 days. Therefore, the raw O’Brien et al. antibody concentration data was used from 0-168 days and the predicted concentrations from Herman et al. were used for the remaining interval, 168-240 days after treatment. This is indicated in Figure 1b by a different line type (solid for *in vivo* concentration data from O’Brien et al. and dashed for modelled concentration data from Herman et al.). In Figure 1d, the dashed line for the concentration data also denotes modelled concentration data rather than *in vivo* measurements of concentration.

### Meta-analysis of monoclonal antibody IC50 and neutralization titer for early pandemic convalescent sera

A previous systematic review and meta-analysis of *in vitro* studies of monoclonal antibodies IC50 against ancestral SARS-CoV-2 and BA.1, BA.2 and BA.1.1 Omicron subvariants has been reported^18^, and provided the raw data from this review. In addition, we have previously extended this systematic review to include studies of *in vitro* IC50 against the BA.4/5 subvariants^12^. Thus, using this previously developed dataset of *in vitro* IC50’s we modified a previously performed meta-analysis of the *in vitro* IC50 of antibodies (bamlanivimab, casirivimab, cilgavimab, imdevimab, sotrovimab, tixagevimab, etesevimab, regdanvimab and bebtelovimab, as these were the antibodies of interest in our previous analysis^12^) to include adintrevimab, as well as the tixagevimab/cilgavimab and casirivimab/imdevimab combinations (Table S2). This meta-analysis involved performing a mixed-effects linear regression (with censoring if IC50 above 10,000) using the *lmec* package^39,40^, as described previously^12^. From this meta-regression we derive a central estimate of the IC50 for each antibody-variant combination (casirivimab/imdevimab, tixagevimab/cilgavimab, and adintrevimab, see Figure S1) considered here, which we use to calculate the *in vivo* antibody concentration as a fold of the *in vitro* IC50 as described below. In addition, we searched all studies included in the systematic review described above for additional data on whether the study reported neutralizing antibody titers for a panel of convalescent subjects from early in the pandemic. We identified 11 studies in which a panel of convalescent sera (obtained from individuals with infections that occurred early in the pandemic, when ancestral viral linages dominated transmission) were assessed for their neutralizing titers^41-51^. Additionally, we identified a further 2 studies where a panel of convalescent sera from individuals infected pre-delta variant of concern were assessed^52,53^. We included the (inverse-transformed) geometric mean neutralization titers reported for these panels of convalescent sera in our meta-regression in order to estimate a mean neutralization titer of early pandemic convalescent sera across the same set of studies as was used to estimate a mean IC50 (Figure S1). This mean convalescent titer across the studies was used to estimate the neutralization titer (on the fold-convalescent scale) for each monoclonal antibody based on the concentration data reported in each prophylaxis study as is described below.

### Estimation of antibody concentration on fold-IC50 scale and estimate of neutralization titer on fold-convalescent scale

Antibody concentrations in each study were extracted and normalized as a ratio to the IC50 from the meta-regression described above (Table S2, reported as antibody concentration, fold-IC50). Note that since some of these studies used antibody combination therapies, the total antibody concentration (sum of both antibody components) was used for the antibody concentration. When comparing the efficacy of these monoclonal antibody trials against that seen after vaccination, we used the estimated neutralization titer in individuals treated with mAbs, converted to a fold-of-convalescent scale since all vaccine trials has been aligned based on this scale previously^14^. This was estimated by taking the antibody concentration as a fold of the IC50 described here, and dividing by the mean convalescent neutralization titer (as described previously^12^).

### Test of concentration effect on the efficacy of prophylactic mAb treatment

Since the concentration data is very sparse below a concentration of 1,000-fold *in vitro* IC50, we tested if there is a significant effect of the antibody concentration on the efficacy in the data. For this analysis, we used a log-binomial regression model, a generalised linear mixed model (GLMM) with a binomial error family and logarithmic link function (using the *glmer* function of the *lme4* package^54^ in R version 4.2.1^55^). The model includes random intercepts for different trials, the covariate “treatment” and the interaction of treatment with concentration. The significance of the interaction of treatment and concentration was tested with a chi-squared test (with the function *drop1*).

### Dose-response fitting with maximum likelihood approach

To estimate the dose-response curve, we fitted a logistic function to the efficacy by concentration data (Figure 2, Figure 4). As we aimed to estimate the concentration that gives 50% protection, we scaled the logistic function such that this concentration is directly estimated as a parameter of the function. The efficacy for a treatment with mAb concentration *c* is then given by

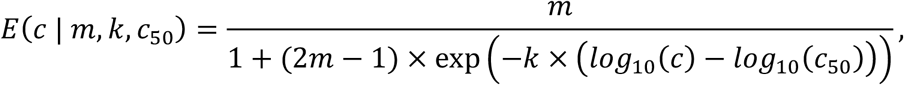

where *m* denotes the maximal efficacy, *k* is a slope parameter and *c*_50_ is the concentration that gives 50% efficacy. Note that this rescaled logistic function will be ill-defined when *m* is below 50%.

We fitted this dose-response curve to the data with a maximum likelihood approach (as previously described^12^). Briefly, the likelihood function used in this optimisation was,

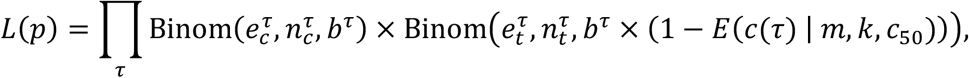

where *p* denotes the parameters of the likelihood function, i.e. the three parameters of the efficacy function (*m*, *k*, *c*_50_) and the baseline risk *b*^*τ*^ for each trial/time interval combination (*τ*), Binom is the probability mass function of the binomial distribution, and for each trial/time interval combination, 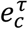 and 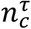 are the numbers of events (symptomatic infections) and total number of individuals in the control group, 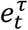 and 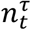 are the numbers of events and total number of individuals in the treatment group in the corresponding trial time interval, and *b*^*τ*^ is the baseline risk which is reduced by the efficacy of treatment for the treatment group in the same interval. The initial guess of the trial/time interval combinations baseline risk used in the optimization were 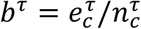. The parameter *c*(*τ*) is the (log10) concentration of monoclonal antibodies (in the fold IC50 scale) in the trial time interval *τ*.

The negative log-transform of this likelihood function was minimised using the *nlm* optimiser in the R statistical software package to estimate the log-transform of the model parameter *k*, *c*_50_ and *m*. The optimiser was run 25 times using randomly generated initial parameters for the transformed parameters *m*, *k*, *c*_50_ drawn uniformly from the ranges log(*m*) = [log(0.6) , log(0.99)], log(*k*) = [log(0.1) , log(100)] and log(*c*_50_) = [log(10) , log(5000)].

Model fitting was used for parameter estimation and hypothesis testing, the latter using a likelihood ratio test of nested models, and Akaike Information Criterion (AIC) for non-nested models. In all fitting, we excluded the earliest time interval of each study to account for the rapid change of the antibody concentration over this time interval and ensured exclusion of treatments which occurred after an unidentified infection (see Table S1 for the time intervals included in the analysis).

The confidence intervals of the fitted model and the fitted model parameters were estimated by parametric bootstrapping. The covariance matrix of the fitted parameters was calculated from the inverse of the Hessian from the model fit^21^. The log-transform of the three model parameters *m*, *k*, and *c*_50_ were drawn randomly from a multinomial Gaussian distribution 10,000 times, using the covariance matrix (and the function *rmvnorm* from the package *mvtnorm* in the R statistic package^56^). The 2.5% and 97.5% percentiles of the estimated parameters provided an estimate of the 95% confidence intervals. Similarly, evaluating the confidence intervals of the fitted model in Figure 2 was estimated by evaluating the model at each neutralisation titre using each of the 10,000 bootstrapped parameter estimates and taking the 2.5% and 97.5% percentiles of the evaluated models at each antibody concentration.

### Timing of protection

Using the dose-response curve, we can predict the protection over time and how long the protection remains above 50% protection (Figure 3). With the meta-analysis of IC50’s against different variants, the timing of protection can be predicted not only for the ancestral strain but also for variants (Figure 3).

We assumed that the concentration of mAbs declines exponentially from the time of the peak concentration for each antibody (extracted from the data) with a half-life fitted to the data (linear fit to log-transformed concentration, see supplementary methods). With the concentration over time and the efficacy by concentration (Figure 2), we computed the protection over time. For variants, we scaled the concentration data by dividing the fold-IC50 concentration from the meta-analysis by the fold-increase in the IC50 and computed the protective efficacy in the same way as against ancestral. The time from treatment to 50% protection is then the time until the concentration falls to the concentration that gives 50% protection which we computed from the concentration over time (using a linear fit to the log-transformed concentration data, Figure S2). The uncertainty in the time to 50% protection is due to the uncertainty in the concentration that gives 50% protection. The upper and lower bounds for the time to 50% protection are the time to reach the upper or lower bound of the 95% CI of the concentration that gives 50% protection, respectively.

### Comparison of the relationship between neutralizing antibodies and protection for vaccination and monoclonal antibody prophylaxis

To compare the mAb data with the data from vaccine studies (Figure 4a), we aimed to match the mAb data as closely as possible with the vaccine studies. Thus, we restricted the mAb data to 2-3 months after treatment, patients who are PCR-negative at baseline, and cases later than 1-2 weeks after treatment (the start of follow-up in vaccine studies) (Figure 4a). We then used a generalised linear mixed model (GLMM) with a binomial error family and logarithmic link function (see above). The model included random intercepts for different trials and a treatment variable with the factors “control”, “mAb” and “vaccine”.

The treatment effect of mAbs and vaccination was compared by testing if there is a significant difference between the coefficients for mAb treatment and vaccination (using the *glht* function from the *multcomp* package^57^).

To further compare the efficacy of vaccination and prophylactic mAb treatment, we normalized the concentration to a common ‘fold-convalescent’-scale (as previously described^12,14^). Using a maximum likelihood approach (see above), we fitted logistic dose-response curves to the monoclonal antibody and vaccine data (Figure 4). We tested whether there is a significant difference between the monoclonal antibody prophylaxis and vaccination by fitting all data with the same parameters for the two types of treatment (Figure 4b, black curve) and compared this fit to fits which have different parameters for antibody treatment and vaccination (e.g., Figure 4b red and blue curves). Different models were compared with a likelihood ratio test. The p-values for different model comparisons are reported in Figure S3 and parameter values for different fits in Table S6. Confidence intervals were calculated using bootstrapping, as described above.

## Data Availability

All data was extracted from publicly available sources and all extracted data and code will be made available on GitHub upon publication.

## Ethics statement

This work was approved under the UNSW Sydney Human Research Ethics Committee (approval HC200242).

## Funding statement

This work is supported by an Australian government Medical Research Future Fund awards GNT2002073 (to MPD, SJK), MRF2005544 (to SJK and MPD), MRF2005760 (to MPD), an NHMRC program grant GNT1149990 (SJK and MPD), and the Victorian Government (SJK). DSK and SJK are supported by NHMRC fellowships. DC, ZKM and EMW and MPD are supported by NHMRC Investigator grants. KLC is supported by PhD scholarship from the Leukaemia Foundation, the Haematology Society of Australia and New Zealand (HSANZ) and Monash University. The Melbourne WHO Collaborating Centre for Reference and Research on Influenza is supported by the Australian Government Department of Health.

## Competing Interests statement

The authors declare no competing interests.

## Data Availability Statement

All data and code will be made available on GitHub upon publication.

## Notes

### Competing Interest Statement

The authors have declared no competing interest.

### Author Declarations

This study involves the meta-analysis of human data that was all openly available prior to the commencement of the study. We searched MEDLINE, PubMed, Embase and the Cochrane COVID Study Register for randomized control trials of monoclonal antibodies for the prevention of COVID-19. We identified six studies. We included only the five studies where monoclonal antibody concentration as well as clinical outcomes were reported for the same cohort. Data was extracted from all eligible studies and which are listed below: 1.Isa, F., et al. Repeat subcutaneous administration of casirivimab and imdevimab in adults is well-tolerated and prevents the occurrence of COVID-19. Int J Infect Dis 122, 585-592 (2022). 2.Levin, M.J., et al. Intramuscular AZD7442 (Tixagevimab-Cilgavimab) for Prevention of Covid-19. N Engl J Med 386, 2188-2200 (2022). 3.O'Brien, M.P., et al. Subcutaneous REGEN-COV Antibody Combination to Prevent Covid-19. N Engl J Med 385, 1184-1195 (2021). 4.Herman, G.A., et al. Efficacy and safety of a single dose of casirivimab and imdevimab for the prevention of COVID-19 over an 8-month period: a randomised, double-blind, placebo-controlled trial. The Lancet Infectious Diseases (2022). 5.Schmidt, P., et al. Antibody-mediated protection against symptomatic COVID-19 can be achieved at low serum neutralizing titers. medRxiv, 2022.2010.2018.22281172 (2022).

### Summary of Updates

The meta-analysis now includes an additional mAb randomised control trial for the antibody Adintrevimab. All analysis, figures, text and supplementary materials have been updated accordingly.

