## Supplementary Material for "Monoclonal antibody levels and protection from COVID-19"

### Supplementary Information

Eva Stadler[1], Martin T Burgess[2], Timothy E Schlub[3], Khai Li Chai[4], Zoe K McQuilten[4,5], Erica M Wood[4,5], Mark N Polizzotto[6,7], Stephen J Kent[8,9], Deborah Cromer[1], Miles P Davenport[1#], David S Khoury[1#].

1. Kirby Institute, University of New South Wales, Sydney, Australia
2. School of Mathematics and Statistics, University of New South Wales, Sydney, Australia
3. Sydney School of Public Health, Faculty of Medicine and Health, University of Sydney, Sydney, New South Wales, Australia
4. Transfusion Research Unit, School of Public Health and Preventive Medicine, Monash University, Melbourne, VIC, Australia
5. Department of Clinical Haematology, Monash Health, Clayton, VIC, Australia
6. Clinical Hub for Interventional Research, College of Health and Medicine, The Australian National University, Canberra, ACT, Australia
7. Department of Clinical Haematology, Canberra Region Cancer Centre, The Canberra Hospital, ACT, Australia
8. Department of Microbiology and Immunology, University of Melbourne at the Peter Doherty Institute for Infection and Immunity, Melbourne, VIC, Australia
9. Melbourne Sexual Health Centre and Department of Infectious Diseases, Alfred Hospital and Central Clinical School, Monash University, Melbourne, VIC, Australia

**# corresponding authors:**
David Khoury
L6 Wallace Wurth Building,
Kirby Institute, UNSW Sydney
Kensington, NSW 2052


Miles Davenport
L6 Wallace Wurth Building,
Kirby Institute, UNSW Sydney
Kensington, NSW 2052


### Supplementary Methods

#### Extraction of case data and efficacy with 95% confidence bounds

We extracted data from the four studies of the number of cases in treatment and control groups at different time intervals after administration of treatment. The source for the case data and the frequency and duration of time intervals for which cases could be extracted are included in the overview of studies (**Table S1**). Data was extracted from figures using WebPlotDigitizer[1] or Adobe Illustrator[2].

For the studies by Isa et al.[3] and Levin et al.[4], the number of patients at risk in the different reported time intervals in the placebo and the treatment group could be extracted along with the number of cases (symptomatic infections). In the studies by O'Brien et al.[5] and Herman et al.[6], the number of patients at risk in the different time intervals could not be extracted. For these studies, we assumed that the number of patients at risk reduced by the number of events in previous time intervals, i.e. we assumed the effect of patients being lost to follow-up is negligible compared to case numbers.

For the study by Schmidt et al.[7], the number of patients at risk was not provided. We used the number of symptomatic infections and percentage of the patients at risk to estimate the number of patients at risk (rounded to the nearest integer).

We computed the efficacy and confidence intervals at each time interval (**Table S1**) from the number of events and patients at risk for the treatment and control group in that interval. Efficacy was estimated as 1 – relative risk (as reported previously[8]), i.e.

$$1-\frac{\text{number events in treatment group/number of patients in treatment group}}{\text{number events in control group/number of patients in control group}}.$$

The 95% confidence intervals for the efficacy estimates were computed using the Katz-log method specified in supplementary table 1 of Aho & Bowyer[9] (as previously[10]). Thus, the 95% confidence intervals for the efficacy are

$$\begin{cases}\left[1-\dfrac{e_t/n_t}{e_c/n_c}\times\exp\left(\pm 1.96\times\sqrt{\dfrac{1}{e_t}+\dfrac{1}{e_c}-\dfrac{1}{n_t}-\dfrac{1}{n_c}}\right)\right], & \text{if } e_t>0 \text{ and } e_c>0,\\ \left[1-\dfrac{(e_t+0.5)/n_t}{e_c/n_c}\times\exp\left(1.96\times\sqrt{\dfrac{1}{e_t+0.5}+\dfrac{1}{e_c}-\dfrac{1}{n_t}-\dfrac{1}{n_c}}\right),1\right], & \text{if } e_t=0,\\ \left[1-\dfrac{e_t/n_t}{(e_c+0.5)/n_c}\times\exp\left(\pm 1.96\times\sqrt{\dfrac{1}{e_t}+\dfrac{1}{e_c+0.5}-\dfrac{1}{n_t}-\dfrac{1}{n_c}}\right)\right], & \text{if } e_c=0,\end{cases}$$

where $e_t$ and $e_c$ are the numbers of events (symptomatic infections) in the treatment and control groups respectively and $n_t$ and $n_c$ are the numbers of patients at risk in the treatment and the control group respectively. If this expression is negative, then the lower bound is set to 0. If there are 0 events in both the treatment and control group, then the efficacy and confidence intervals are not defined at that time interval.

**Extraction of concentration data and estimating the geometric mean concentration for given time intervals**

We also extracted concentration data from the different studies (**Table S1**) using WebPlotDigitizer[1] or Adobe Illustrator[2]. For the studies by Isa et al.[3], Herman et al.[6] and Levin et al.[4], we extracted an estimate of the total concentration of both antibodies used in combination (i.e. casirivimab + imdevimab or tixagevimab + cilgavimab, respectively). For the study by O'Brien et al.[5] we extracted the concentration of casirivimab and imdevimab separately and added these concentrations together to estimate the total concentration of both casirivimab and imdevimab. For the study by Schmidt et al.[7], we extracted the modelled concentration data, since the raw concentration data were visually obscured due to the high number of overlapping concentration data points.

We estimated the geometric mean concentration for each time interval over which efficacy data was available by first linearly interpolating between the concentrations reported at each time point in the extracted concentration data (using the function *approx* in R[11]). The geometric mean concentration for each time interval was computed as,

$$\exp\left(\frac{\int_{t_{min}}^{t_{max}} \log(c(t))\, dt}{t_{max} - t_{min}}\right),$$

where *c(t)* is the linearly interpolated concentration by time and $t_{min}$ and $t_{max}$ are the lower and upper end of the time interval. The geometric mean concentration was computed for each study and each time interval of case data (see horizontal error bars in **Figure 1**).

**Estimating antibody half-life**

In order to predict the duration of protection of each monoclonal antibody combination, we first estimated the *in vivo* half-life for casirivimab/imdevimab, tixagevimab/cilgavimab and adintrevimab. The half-lives were estimated by fitting a linear regression model to the log-transformed concentration data from the time of the peak antibody concentration onwards. This fitting was performed using the *lm* function in R and the function *confint* to determine 95% confidence intervals for the estimated half-lives. We found that the estimated half-lives agree well with the mean of the half-lives of casirivimab and imdevimab or tixagevimab and cilgavimab that were reported in the literature (**Table S6**).

### Supplementary Tables

**Overview of prophylactic mAb studies**

| Trial with reference | mAb | Dose [mg] | Admin. | Notes/Study description | Case data source | Duration of follow-up used in analysis (days post administration) | Time intervals for which cases could be extracted (days post administration) | Antibody concentration data source |
|---|---|---|---|---|---|---|---|---|
| **Isa[3]** | casirivimab/ imdevimab | 1,200 | SC | Administered up to 6 doses every 4 weeks | Figure 1 | 28 – 167 | 1-27*, 28-55, 56-83, 84-111, 112-139*, 140-167, 168-196* | Figure 2 |
| **O'Brien[5]** | casirivimab/ imdevimab | 1,200 | SC | Treatment of household contacts of confirmed cases Subgroup of seronegative at baseline used here | Table S4 | 8 – 28 | 1-7$^{\lozenge}$, 8-14, 15-21, 22-28 | Figure S4 |
| **Herman[6]** | casirivimab/ imdevimab | 1,200 | SC | Same as O'Brien study but with extended follow-up (8 months) | Figure 2A | 31 – 240 | 1-30$^{\lozenge}$, 31-60, 61-90, 91-120, 121-150, 151-180, 181-210, 211-240 | Table S6 |
| **Levin[4] (PROVENT)** | tixagevimab/ cilgavimab | 300 | IM | Treatment of high risk patients (inadequate response to vaccination and/or high exposure) | Figure 2 | 11 – 183 | 1-10$^{\#}$, 11-29, 30-59, 60-89, 90-119, 120-149, 150-179, 180-183 | Figure S2A |
| **Schmidt[7]** | adintrevimab | 300 | IM | Phase 2/3 clinical study of efficacy during the Delta and Omicron BA.1/BA.1.1 waves | Table S3 | Pre-Omicron: 91 – 180<br>Omicron: 29 – 90 | Pre-Omicron: 0-90$^{\lozenge}$, 91-180<br>Omicron: 1-28$^{\lozenge}$, 29-60, 61-90 | Figure 2A (model) |
| **Cohen[12] (BLAZE-2)** | bamlanivimab | 4,200 | IV | Treatment of residents or staff in a skilled nursing or assisted living facility with at least one confirmed case of SARS-CoV-2 ≤7 days prior to randomization | Not included in analysis | Not included in analysis | - | No Concentration data |

**Table S1** Overview over identified prophylactic mAb studies. The earliest cases were excluded for all studies (note that for the Isa study there were no cases in either the treatment or the control group in the first 28 days after treatment). Abbreviations: SC subcutaneous, IM intramuscular, IV intravenous. * For these time intervals, there were no cases in both the treatment and the control group, thus the efficacy could not be computed, and the time intervals were excluded. $^{\#}$ For the earliest time interval in the study by Levin et al., the geometric mean concentration could not be computed as the first data point for the *in vivo* concentration is on day 8. We excluded this time interval from the analysis. $^{\lozenge}$ The earliest time intervals in the O'Brien et al., Herman et al. and Schmidt et al. studies are shown in **Figure 2** but were excluded from the analysis (see Methods).

**Estimated *in vitro* IC50 for monoclonal antibodies against ancestral virus and Omicron subvariants from systematic review and meta-analysis**

| Antibodies | Ancestral | BA.1 | BA.1.1 | BA.2 | BA.4/5 |
|---|---|---|---|---|---|
| Adintrevimab | 6.55 (95% CI: 3.63 - 11.8) | 1360 (95% CI: 635 - 2900) | 702 (95% CI: 296 - 1670) | >10000 | >10000 |
| Bamlanivimab | 6.01 (95% CI: 3.98 - 9.09) | >10000 | >10000 | >10000 | >10000 |
| Bebtelovimab | 3.31 (95% CI: 1.76 - 6.25) | 4.15 (95% CI: 2.02 - 8.56) | 2.71 (95% CI: 0.952 - 7.74) | 4.28 (95% CI: 1.8 - 10.31) | 4.84 (95% CI: 1.46 - 16.1) |
| Casirivimab | 5.17 (95% CI: 3.47 - 7.7) | >10000 | >10000 | >10000 | >10000 |
| Casirivimab/ Imdevimab | 3.26 (95% CI: 1.92 - 5.53) | >10000 | >10000 | 2530 (95% CI: 1030 - 6190) | 4510 (95% CI: 1270 - >10000) |
| Cilgavimab | 8.05 (95% CI: 5.37 - 12.1) | 5310 (95% CI: 3260 - 8630) | >10000 | 25.6 (95% CI: 13.6 - 48.2) | 93.3 (95% CI: 32.8 - 266) |
| Cilgavimab/ Tixagevimab | 4.00 (95% CI: 2.44 - 6.57) | 262 (95% CI: 139 - 493) | 1640 (95% CI: 491 - 5480) | 38.4 (95% CI: 17.1 - 86.2) | 727 (95% CI: 254 - 2090) |
| Etesevimab | 23.4 (95% CI: 15.5 - 35.4) | >10000 | >10000 | >10000 | >10000 |
| Imdevimab | 8.77 (95% CI: 5.94 – 12.9) | >10000 | >10000 | 1780 (95% CI: 887 - 3570) | 9090 (95% CI: 2900 - >10000) |
| Regdanvimab | 2.29 (95% CI: 1.16 - 4.54) | >10000 | >10000 | >10000 | >10000 |
| Sotrovimab | 81.8 (95% CI: 55.8 - 120) | 256 (95% CI: 162 - 406) | 289 (95% CI: 122 - 685) | 1370 (95% CI: 722 - 2590) | 2430 (95% CI: 855 - 6940) |
| Tixagevimab | 3.11 (95% CI: 2.08 - 4.64) | 2100 (95% CI: 1370 - 3220) | 4010 (95% CI: 1640 - 9800) | 6380 (95% CI: 2950 - >10000) | 7520 (95% CI: 2330 - >10000) |
| Convalescent subjects | 618* (95% CI: 335 – 1140) | NA | NA | NA | NA |

**Table S2** The geometric mean IC50 (in ng/mL) for monoclonal antibodies estimated using a meta-analysis of data from the systematic review[13] (plus extended data including BA.4/5 from Yamasoba et al.[14], Tuekprakhon et al.[15], Arora et al.[16] and Wang et al.[17]) for ancestral and select Omicron subvariants. The meta-regression was performed on the log-transformed IC50 values reported for each antibody variant combination using a linear mixed effects model with a random effect for study and censoring of IC50's above 10,000. Where the estimated geometric mean IC50 was above 10,000 the value reported is ">10,000". *Where studies in the systematic review[13] included a panel of convalescent serum assessed for neutralization in the same assay as that used for the IC50, the mean titer reported by the study was extracted and the inverse of this titer was included in the meta-regression. Here we reported the mean neutralization titer estimated across studies for convalescent subjects. Highlighted rows in grey were used to estimate the antibody concentration on a fold IC50 scale (e.g. **Figure 2**), and the neutralization antibodies in subjects after administration on a fold-convalescent scale (**Figure 4**, **Figure S5**).

**Relationship between efficacy and mAb concentration (leave-one-out analysis)**

| Data | Chi-squared test p-value |
|---|---|
| All | $9 \times 10^{-10}$ |
| All but O'Brien et al. study | $2 \times 10^{-7}$ |
| All but Herman et al. study | $1 \times 10^{-8}$ |
| All but Isa et al. study | $4 \times 10^{-8}$ |
| All but Levin et al. study | $2 \times 10^{-8}$ |
| All but Schmidt et al. study (delta) | $1 \times 10^{-9}$ |
| All but Schmidt et al. study (omicron) | 0.011 |
| All but Schmidt et al. study (either variant) | 0.016 |

**Table S3** Assessing whether there was a significant association between the efficacy and the mAb concentration using a generalized linear mixed effects model with a binomial error family and logarithmic link function, and a chi-squared test for the significance of the mAb concentration as a covariate (see Methods). There is a significant relationship between the mAb concentration and the efficacy in the data, even if one study is excluded.

**Parameter estimates for the dose-response curve**

| Parameter | Estimate | 95% confidence interval |
|---|---|---|
| Maximal efficacy | 94.4% | 83.9 – 106.3% |
| Slope parameter | 1.3 | 0.6 – 2.9 |
| Concentration that gives 50% protection [fold *in vitro* IC50] | 54.3 | 16.1 – 183.0 |

**Table S4** Parameter estimates and 95% confidence intervals for the dose-response curve of efficacy by concentration [fold *in vitro* IC50]. These parameters were estimated by model fitting (**Figure 2**).

**Days of >50% protection against variants**

| | Ancestral | BA.1 | BA.1.1 | BA.2 | BA.4/5 |
|---|---|---|---|---|---|
| **Adintrevimab** | 903 (667 – 1139) | 0 (0 – 103) | 0 (0 – 231) | 0 (0 – 0) | 0 (0 – 0) |
| **Casirivimab/ imdevimab** | 269 (219 – 320) | 0 (0 – 0) | 0 (0 – 0) | 0 (0 – 44) | 0 (0 – 20) |
| **Tixagevimab/ cilgavimab** | 670 (503 – 836) | 97 (0 – 263) | 0 (0 – 0) | 360 (194 – 526) | 0 (0 – 124) |

**Table S5** Duration of protection against symptomatic infection of at least 50% with 95% confidence interval for different antibody and variant combinations. The duration of protection in days was computed using the time of peak concentration and the half-life fitted to the concentration data for each mAb (**Table S6**) and the best fit efficacy model to the mAb data (**Figure 2** and **Table S4**).

**Temporal kinetics of casirivimab/imdevimab and tixagevimab/cilgavimab concentrations**

| | **Adintrevimab** | **Casirivimab/imdevimab** | **Tixagevimab/cilgavimab** |
|---|---|---|---|
| Source of concentration data | Schmidt | O'Brien & Herman | Levin |
| Peak concentration [mg/L] | 36.3 | 108.1 | 23.9 |
| Day of peak concentration | 6 | 3 | 29 |
| Half-life estimated from the data [days] | 134.7 | 28.8 | 94.9 |
| 95% CI of the estimated half-life | 132.7 – 136.7 | 26.6 – 31.3 | 84.2 – 108.9 |
| Half-life from the literature [days] | 141[7] | 29.8[18] | 86.5[19] |

**Table S6** Summary of the kinetics of the antibody concentration over time for casirivimab/imdevimab and tixagevimab/ cilgavimab. For casirivimab/imdevimab, only the data from the O'Brien and Herman studies was used, since individuals were re-treated monthly in the Isa study. The half-life from the data was estimated by fitting a linear model to the log-transformed concentration data from the peak. The half-life from the literature is the mean of the reported half-lives of casirivimab and imdevimab or tixagevimab and cilgavimab, respectively. Abbreviation: CI confidence interval.

**Protective efficacy of the peak mAb concentration against variants**

| | **Adintrevimab** | **Casirivimab/imdevimab** | **Tixagevimab/cilgavimab** |
|---|---|---|---|
| **Ancestral** | 88.5% (76.9 – 92.9) | 92.2% (82.5 – 96.6) | 88.8% (77.3 – 93.0) |
| **Omicron BA.1** | 40.7% (17.9 – 55.9) | 17.4% (2.0 – 36.2) | 56.7% (38.9 – 71.4) |
| **Omicron BA.1.1** | 49.3% (29.0 – 64.5) | 3.6% (0.0 – 19.8) | 33.1% (10.5 – 49.0) |
| **Omicron BA.2** | 2.4% (0.0 – 16.9) | 46.8% (25.5 – 61.9) | 77.0% (62.8 – 85.4) |
| **Omicron BA.4/5** | 2.0% (0.0 – 15.7) | 39.3% (16.4 – 54.5) | 43.4% (21.1 – 58.5) |

**Table S7** Protective efficacy of prophylactic mAb treatment at peak concentration for different antibody and variant combinations. The peak concentration for each mAb was extracted from the data (**Table S6**) and the best fit efficacy model was used to predict the efficacy and 95% CI of efficacy.

**Fold-changes for >50% protection for >30 days and for variants**

| | **Max fold-change for >50% protection for >30 days (95% CI)** | **Fold-change from ancestral to Omicron** | | |
|---|---|---|---|---|
| | | **BA.1** | **BA.2** | **BA.4/5** |
| **Adintrevimab** | 89.6 (26.6 – 302.0) | 207.0 | 84074.0 | 119941.6 |
| **Casirivimab/imdevimab** | 319.5 (94.8 – 1076.5) | 10796.9 | 776.5 | 1385.5 |
| **Tixagevimab/cilgavimab** | 106.8 (31.7 – 360.0) | 65.5 | 9.6 | 181.8 |

**Table S8** Maximal fold-change of the IC50 of a variant such that the protective efficacy remains above 50% for at least 30 days after treatment and the fold-change of *in vitro* IC50s of Omicron variants BA.1, BA.2 and BA.4/5.

**Parameter values for different model fits comparing the efficacy of vaccination and prophylactic mAbs**

| Model | | m | k | $c_{50}$ | Negative log-likelihood | AIC |
|---|---|---|---|---|---|---|
| same m, k, $c_{50}$ | | 95.27% | 2.64 | 0.20 | 130.1 | 330.3 |
| different m<br>same k, $c_{50}$ | vaccine data<br>mAb data | 103.52%<br>92.31% | 1.86 | 0.17 | 124.9 | 321.9 |
| different k<br>same m, $c_{50}$ | vaccine data<br>mAb data | 98.91% | 2.55<br>1.16 | 0.21 | 124.0 | 320.0 |
| different $c_{50}$<br>same m, k | vaccine data<br>mAb data | 94.41% | 2.86 | 0.21<br>0.09 | 129.9 | 331.8 |
| different m, k<br>same $c_{50}$ | vaccine data<br>mAb data | 99.34%<br>96.41% | 2.47<br>1.31 | 0.21 | 123.9 | 321.8 |
| different m, $c_{50}$<br>same k | vaccine data<br>mAb data | 101.39%<br>90.38% | 2.15 | 0.19<br>0.08 | 124.2 | 322.3 |
| different $c_{50}$, k<br>same m | vaccine data<br>mAb data | 98.23% | 2.76<br>1.04 | 0.23<br>0.09 | 123.1 | 320.2 |
| different m, k, $c_{50}$ | vaccine data<br>mAb data | 98.68%<br>94.43% | 2.68<br>1.29 | 0.22<br>0.09 | 123.0 | 321.9 |

**Table S9** Parameter values for the different model fits to compare vaccine and mAb data. The models were fit using a maximum likelihood approach and a likelihood ratio test was used to compare models (see Methods). Fitted models and comparisons are shown in **Figure 4b**, **Figure S3** and **Figure S5**. If there are two values in a cell for a model, the upper one is the estimate for the vaccine data and the lower is the estimate for the mAb data. If there is only one value, then the vaccine and mAb data estimates are the same. The concentration that gives 50% protection ($c_{50}$) is given in fold-convalescent here (to compare the vaccine and the mAb data the concentrations were transformed to fold-convalescent by dividing by the mean convalescent neutralization titer). Abbreviations: m maximal efficacy, k slope parameter, $c_{50}$ concentration that gives 50% protection.

### Supplementary Figures

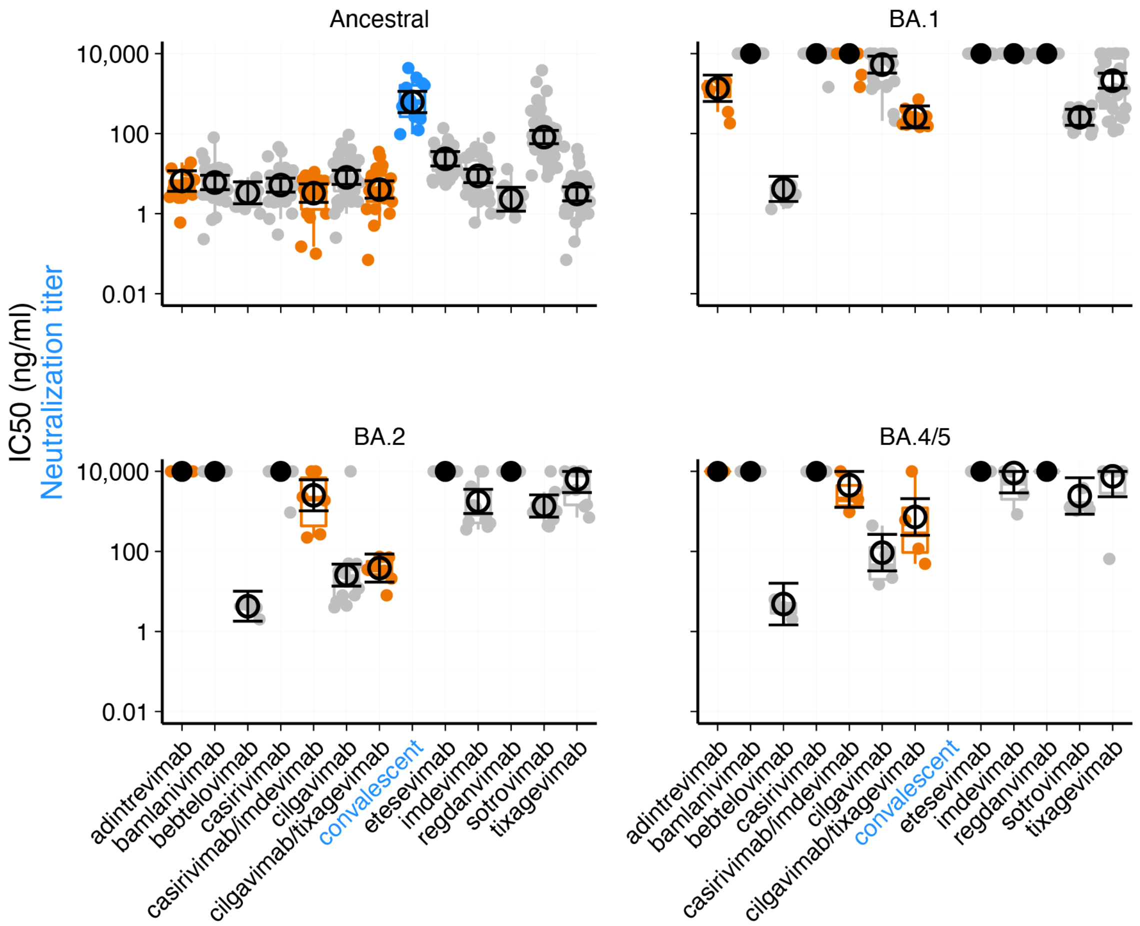


**Figure S1** A boxplot of the IC50 values reported for each antibody (grey and orange) against each variant in each study of the systematic review[13] (with additional data as described in the methods). Each small dot represents the reported IC50 from an individual study and the large open circles are the estimated geometric mean IC50 from the meta-regression (described in the methods). Closed circles indicate that the geometric mean IC50 was above 10,000 ng/ml. The geometric mean neutralization titer of serum collected from a cohort of convalescent individuals was reported in 13 studies from the systematic review, and these are shown (blue) along with the geometric mean neutralization titer from the meta-regression. Orange indicates the antibody combinations that are used in the clinical studies that are the focus of the analysis presented in this study.

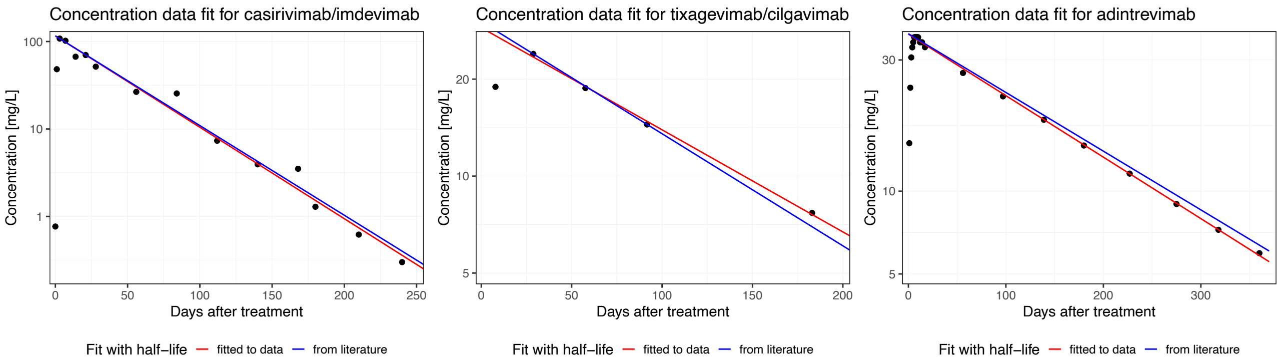


**Figure S2** Antibody concentration data extracted from O'Brien et al.[5] and Herman et al.[6] (left), Levin et al.[4] (middle) and Schmidt et al.[7] (right) for casirivimab/imdevimab, tixagevimab/cilgavimab and adintrevimab, respectively (black dots). Also shown are the best fitting line (linear regression) to the data from the peak reported concentration to the end of the time series (red). The half-life for each antibody combination reported in the literature is shown in blue for comparison (see also **Table S6**).

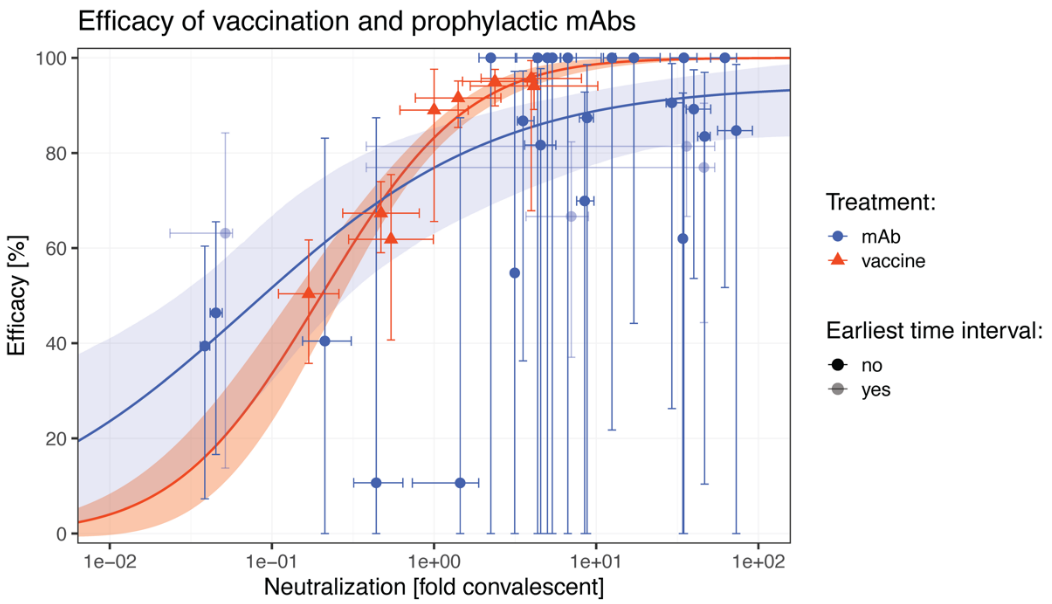


**Figure S3** Comparison of the best fitting efficacy models to the vaccine data (red) and the prophylactic mAb data (blue). The best fitting model to the vaccine data is from Khoury et al.[10] and the best fitting model to the mAb data is the model shown in **Figure 2**.

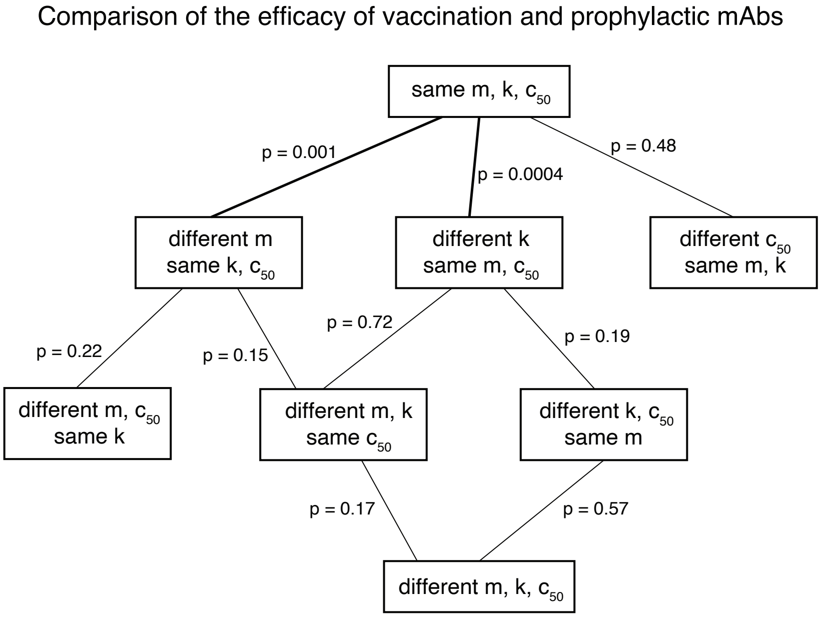


**Figure S4** Comparison of model fits to the vaccination and mAb efficacy data using a forward regression strategy. To compare the efficacy of vaccination and prophylactic mAb treatment, we fitted a logistic dose-response curve to the data (**Figure 4b** and **Figure S5**). The parameters of the dose-response curve are the maximum efficacy (m), a slope parameter (k) and the neutralization titer (on the fold of convalescence scale) that gives 50% protection ($c_{50}$). First, we fitted the dose-response curve to all data and assumed the model parameters are the same for vaccines and mAbs ("same m, k, c50", see black curve in **Figure 4b** and **Figure S5**). Next, we systematically allowed a single parameter to differ between the vaccines and mAbs and compared the resulting model with the first model using a likelihood-ratio test (indicated p-values). This showed the biggest improvement by allowing the slope to vary (see red and blue lines in **Figure 4b**). We fitted further models allowing the slope and either the maximum efficacy or the neutralization titer that gives 50% protection to vary between both types of treatment, but neither significantly improved the model. Finally, the model with all parameters differing between vaccines and mAbs did not significantly improve the model. Thus, we found the best fit is the model that allows for different slopes but has the same maximum efficacy and 50% protection neutralization for vaccines and mAbs (red and blue lines in **Figure 4b**). The parameter values for different model fits can be found in **Table S9**.

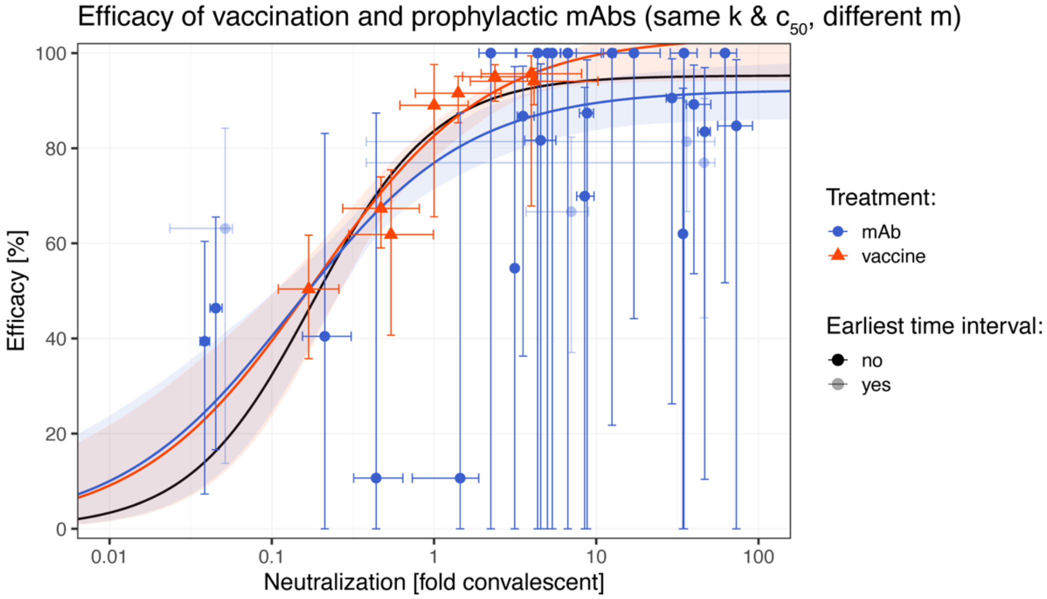


**Figure S5** Comparison of prophylactic mAb treatment and vaccination. The black line is the fit to all data assuming that the maximal efficacy, slope parameter and the neutralization titer that gives 50% protection are the same for the mAbs and vaccines. The colored lines indicate the model fit to this data which allows for different maximal efficacy (m) of mAbs (blue) and vaccines (red) but the same neutralization titer that gives 50% protection ($c_{50}$) and slope parameter. This figure highlights that although we could not detect a significant difference in the maximum efficacy, there is a trend towards a lower protective efficacy for mAbs.
